# Apathy is not associated with reduced ventral striatal volume in patients with schizophrenia

**DOI:** 10.1101/2020.02.07.20019943

**Authors:** Achim Burrer, Fernando Caravaggio, Andrei Manoliu, Eric Plitman, Karoline Gütter, Benedikt Habermeyer, Philipp Stämpfli, Aslan Abivardi, André Schmidt, Stefan Borgwardt, Mallar Chakravarty, Martin Lepage, Alain Dagher, Ariel Graff-Guerrero, Erich Seifritz, Stefan Kaiser, Matthias Kirschner

## Abstract

**Background:** A growing body of neuroimaging research has revealed a relationship between blunted activation of the ventral striatum (VS) and apathy in schizophrenia. In contrast, the association between reduced striatal volume and apathy is less well established, while the relationship between VS function and structure in patients with schizophrenia remains an open question. Here, we aimed to replicate previous structural findings in a larger independent sample and to investigate the relationship between VS hypoactivation and VS volume.

**Methods:** We included brain structural magnetic resonance imaging (MRI) data from 60 patients with schizophrenia (SZ) that had shown an association of VS hypoactivation with apathy during reward anticipation and 58 healthy controls (HC). To improve replicability, we applied analytical methods developed in two previously published studies: Voxel-based morphometry and the Multiple Automatically Generated Templates (MAGeT) algorithm. VS and dorsal striatum (DS) volume were correlated with apathy correcting for age, gender and total brain volume. Additionally, left VS activity was correlated with left VS volume.

**Results:** We failed to replicate the association between apathy and reduced VS volume and did not find a correlation with DS volume. Functional and structural left VS measures exhibited a trend-level correlation (r_s_=0.248, p=0.067, r^2^=0.06).

**Conclusions:** Our present data suggests that functional and structural striatal neuroimaging correlates of apathy can occur independently. Replication of previous findings may have been limited by other factors (medication, illness duration, age) potentially related to striatal volume changes in SZ. Finally, associations between reward-related VS function and structure should be further explored.

## 1. Introduction

Apathy can be defined as a reduction in motivation and goal-oriented behavior and is a core negative symptom of schizophrenia (Brown and Pluck, 2000; Marin, 1996). This debilitating symptom is prevalent in early disease stages (Faerden et al., 2010; Fervaha et al., 2015; Lam et al., 2015), associated with poor treatment compliance (Tattan and Creed, 2001) and is a strong predictor for poor functional outcome and reduced quality of life (Faerden et al., 2013; Fervaha et al., 2018; Galderisi et al., 2013; Strauss et al., 2013).

From a neurobiological perspective it has been observed that functional and structural abnormalities within cortico-striatal circuits are critically involved in the pathophysiology of negative symptoms; in particular apathy and motivational deficits (Ehrlich et al., 2012; Haber, 2016; Hovington and Lepage, 2012; Kos et al., 2016; Li et al., 2018; Radua et al., 2015; van Erp et al., 2016; van Erp et al., 2018; Walton et al., 2018). In schizophrenia, numerous studies demonstrated that severity of apathy is associated with blunted striatal response during reward processing (e.g. reward anticipation, prediction error coding, receipt of reward), with most evidence for a relationship to blunted reward anticipation (Fig. 1) (Arrondo et al., 2015; Dowd and Barch, 2010, 2012; Dowd et al., 2016; Kirschner et al., 2016; Kluge et al., 2018; Moran et al., 2019; Morris et al., 2015; Mucci et al., 2015; Simon et al., 2010; Simon et al., 2015; Stepien et al., 2018; Waltz et al., 2009; Waltz et al., 2010; Waltz et al., 2018; Wolf et al., 2014). While most studies in schizophrenia found reduced blood-oxygen-level-dependent (BOLD) activity related to apathy in the ventral striatum (VS), some studies have identified a specific involvement of the dorsal striatum (DS) (Morris et al., 2015; Mucci et al., 2015), or dysfunctions including both VS and DS (Dowd et al., 2016; Moran et al., 2019; Stepien et al., 2018).

**Figure 1.**
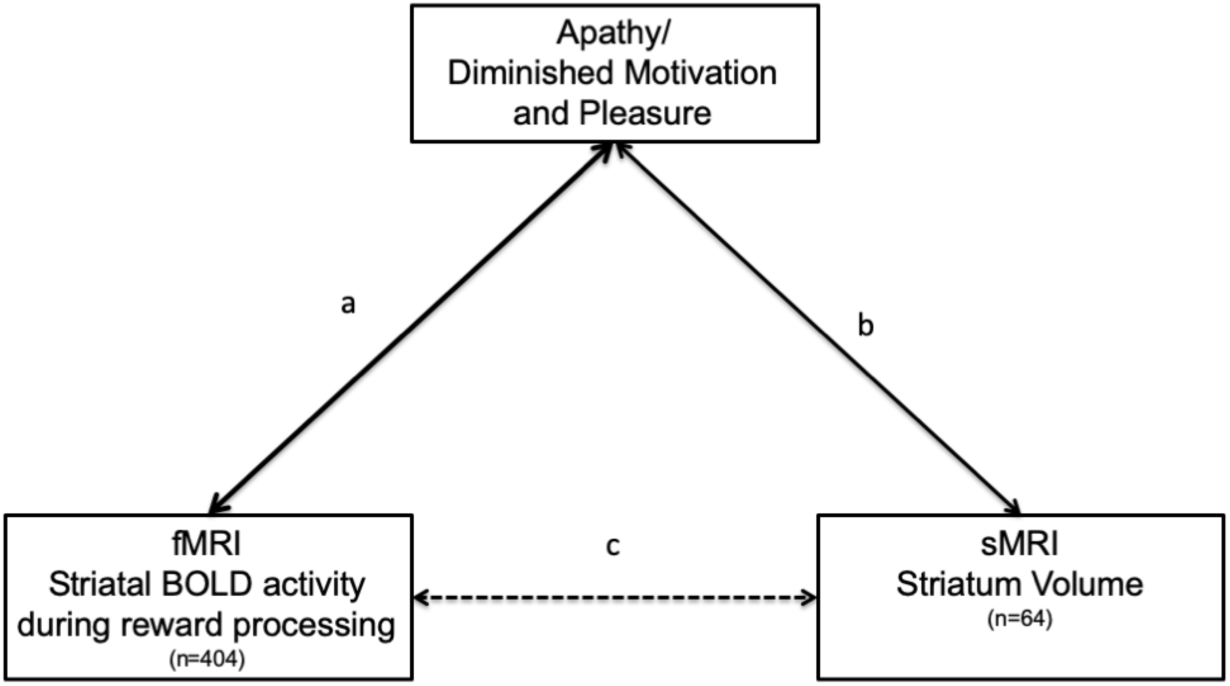
Overview studies investigating striatal correlates of apathy. (a) 14 studies (n=355) reported an association between apathy/diminished motivation and pleasure and striatal activity during processing of reward and salient stimuli (Arrondo et al., 2015; Dowd and Barch, 2010, 2012; Dowd et al., 2016; Kirschner et al., 2016; Kluge et al., 2018; Moran et al., 2019; Morris et al., 2015; Mucci et al., 2015; Simon et al., 2010; Simon et al., 2015; Stepien et al., 2018; Waltz et al., 2009; Waltz et al., 2010; Waltz et al., 2018; Wolf et al., 2014). (b) 2 studies (n=64) reported an association between apathy/diminished motivation and pleasure and striatal volume (Caravaggio et al., 2018a; Roth et al., 2016). (c) Hypothesized relation between VS volume and VS BOLD activity during reward processing in patients with schizophrenia.

Contrasting this wealth of evidence on the relationship between aberrant striatal function and motivational deficits in schizophrenia, potential structural abnormalities of the striatum have received little attention. To our knowledge only three studies have directly investigated the association between apathy and subcortical structural abnormalities. Two studies investigated correlative relationships between VS volume and measures of apathy (Caravaggio et al., 2018a; Roth et al., 2016), while one study compared subcortical volumes (including caudate, putamen and accumbens) between patients with persistent apathy and patients without apathy (Morch-Johnsen et al., 2015). Using Voxel-based morphometry, Roth et al. found that apathy (measured with the Apathy Evaluation Scale) was associated with reduced right VS volume, but not left VS volume, in 23 individuals with a diagnosis of schizophrenia or schizoaffective disorder (Roth et al., 2016). This finding was replicated by Caravaggio and colleagues who observed a relationship between apathy (PANSS amotivation factor) and both left and right VS volume using a different method – the “Multiple Automatically Generated Templates (MAGeT-Brain)” algorithm (Caravaggio et al., 2018a). The study from Caravaggio and colleagues also extended previous work showing in 41 older schizophrenia patients that the relationship between VS volume was specifically related to individual levels of amotivation (not diminished expression) and independent of antipsychotic occupancy of striatal dopamine D2/3 receptors measured with positron emission tomography (Caravaggio et al., 2018a). Contrary to these findings, Morch-Johnsen and colleagues observed a trend level larger caudate volume in a small sample of first episode patients with persistent apathy (n = 18) compared to patients with non-persistent apathy (Morch-Johnsen et al., 2015). In contrast, earlier studies investigating global negative symptoms did not find a correlation with reduced striatal volume (Ehrlich et al., 2012). Taken together, studies on striatal structural abnormalities are inconsistent and there is a dramatic discrepancy between the large body of research focusing on functional striatal correlates compared to the few studies addressing structural correlates of apathy in schizophrenia (Fig. 1). Specifically, it is unclear whether functional and structural VS abnormalities related to apathy occur simultaneously in the same patients and covary with each other. Moreover, no study has directly addressed both correlates of apathy using a multimodal approach combining functional and structural magnetic resonance imaging (fMRI/sMRI) measures of the striatum. In the present study, we aimed to bridge this gap combining structural T1-weighted images from three previously published fMRI studies, investigating the association of VS dysfunction during reward anticipation with apathy in patients with schizophrenia (Kirschner et al., 2016; Kluge et al., 2018; Stepien et al., 2018) (Fig. 2).

**Figure 2.**
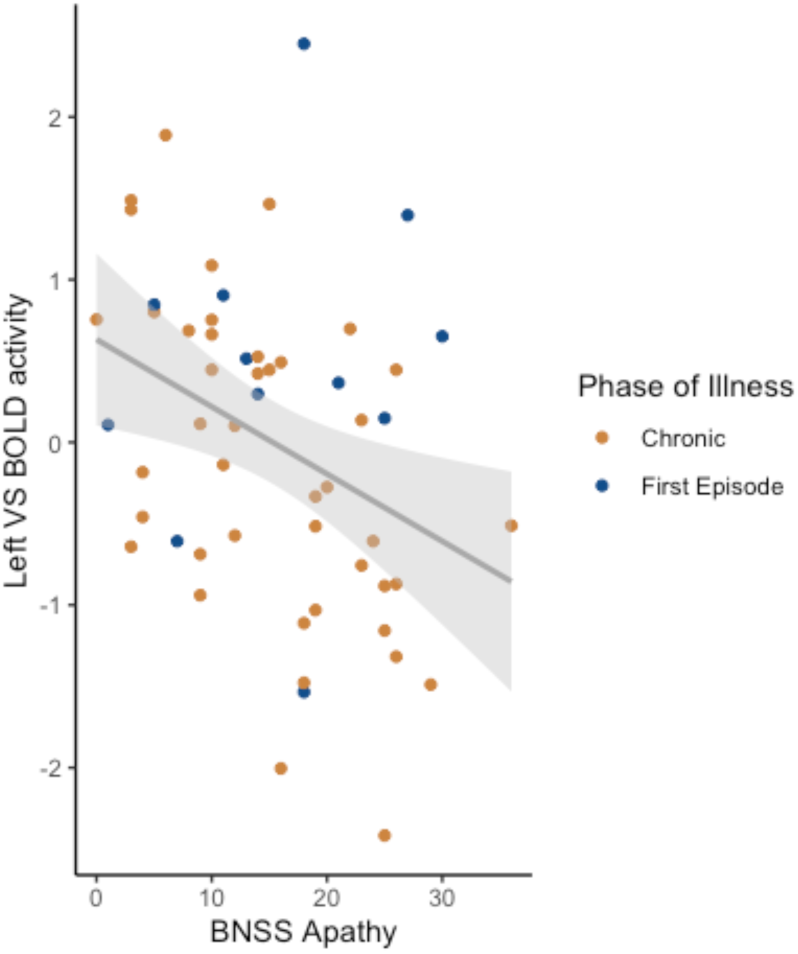
Non-parametric Spearman correlation between apathy and left VS BOLD activity during reward anticipation (r_s_=-.41, p=0.003) across pooled data (n=55) from three previous published publications (Kirschner et al., 2016; Kirschner et al., 2018; Stepien et al., 2018).

First, we asked whether the reported negative association between VS volume and apathy (Caravaggio et al., 2018a; Roth et al., 2016) can be replicated in an independent, larger sample and if this extends to the DS. To improve comparability with these studies, we applied the same analytical approaches: (a) voxel-based-morphometry used by Roth an colleagues (Roth et al., 2016) and (b) the MAGeT Brain algorithm used by Caravaggio and colleagues (Caravaggio et al., 2018a). We hypothesized that apathy would be correlated with reduced VS volume resulting in a simultaneously pattern of both functional and structural VS correlates of apathy (Fig. 1a, b). Second, we asked whether functional and structural abnormalities are correlated with each other (Fig. 1c). Specifically, we hypothesized that blunted VS activation during reward anticipation is associated with reduced VS volume.

## 2. Materials and Methods

### 2.1 Participants

We included data from three previous fMRI studies investigating the relationship between striatal activity during reward anticipation and apathy in patients with schizophrenia (Kirschner et al., 2016; Kirschner et al., 2018; Stepien et al., 2018). In total, 60 patients diagnosed with schizophrenia (SZ) and 58 healthy controls (HC) were included in the present study. Patients with SZ were recruited from inpatient and outpatient units of the Psychiatric Hospital of the University of Zurich. HC were recruited from the general community via advertisement. SZ diagnosis was confirmed using the structured Mini-International Neuropsychiatric Interview for DSM-IV (Lecrubier et al., 1999). Exclusion criteria were: (1) any other DSM-IV Axis I disorder (in particular current substance use disorder and major depressive disorder), (2) benzodiazepine medication higher than 1 mg/d lorazepam equivalent, (3) Psychotic symptoms scoring higher than 4 on the positive subscale of the Positive and Negative Syndrome Scale (PANSS) (Kay et al., 1989) and, (4) extrapyramidal symptoms scoring higher than 2 on the Modified Simpson-Angus Scale (MSAS) (Simpson and Angus, 1970). Patients were clinically stable and had been on a stable dose of medication for at least 2 weeks before testing. HC were screened for any neuropsychiatric disorders using the structured Mini-International Neuropsychiatric Interview (Lecrubier et al., 1999). Both patients and HC were required to have a normal physical and neurological examination and no history of major head injury or neurologic disorder. The Ethics Committee of the Canton of Zurich approved the project and participants gave written informed consent to participate in the study. The capacity of each participant with SZ to provide informed consent was evaluated by the treating psychiatrist.

### 2.2 Clinical and neuropsychological assessment

Negative symptoms were assessed using the Brief Negative Symptom Scale (BNSS) (Kirkpatrick et al., 2011). The apathy dimension score included avolition, asociality and anhedonia, while the diminished expression dimension score included alogia and blunted affect. Additional, psychopathological assessment included the Positive and Negative Symptoms Scale (PANSS) (Kay et al., 1989), the Calgary Depression Scale for Schizophrenia (CDSS) (Addington et al., 1990) and the Global Assessment of Functioning Scale (GAF).

### 2.3 MRI data acquisition

MRI data were acquired on a Philips Achieva 3.0T whole-body scanner (Best, The Netherlands) (upgraded to the dStream platform). A 32-channel received head coil (Philips, Best, The Netherlands) and MultiTransmit parallel radio frequency transmission was used. T1-weigthed images were collected via a 3D magnetization-prepared rapid gradient-echo sequence (MP-RAGE) with the following parameters: acquisition voxel size=1.0×1.0×1.0mm^3^, time between two inversion pulses=2997ms, inversion time=1008ms, inter-echo delay = 8.1ms, flip angle=8°, matrix=240×240, field of view=240×240mm^2^, 160 sagittal slices. Total scan duration was 07:32.0.

### 2.4 Subcortical Volume Analysis

We examined the association between striatal volume and apathy using the same structural measures used in previous studies; namely: voxel-based morphometry (Ashburner and Friston, 2000) and the Multiple Automatically Generated Templates algorithm(Chakravarty et al., 2013; Pipitone et al., 2014).

### 2.5 Voxel-based morphometry (VBM)

We performed voxel-based morphometry (VBM) using the statistical parametric mapping package (SPM 12; Wellcome Trust Centre for Neuroimaging, London) under the MATLAB R2016a platform (MathWorks, Inc., Sherborn, USA). The automated segmentation function was used to separate white matter, grey matter and cerebrospinal fluid. Afterwards the segmented images were normalized to Montreal Neurological Institute (MNI) space using Diffeomorphic Anatomical Registration Through Exponentiated Lie Algebra (DARTEL) and smoothed with a 3mm full-width at half maximum (FWHW) Gaussian Filter. The voxel size of data acquisition was 1×1×1mm^3^.Total grey matter volume (TGMV) was derived from the first segmentation process (Manoliu et al., 2014). We performed quality control of data and pre-processing steps by visual inspection of raw images, grey matter, white matter and CSF segmentation. No scan had to be excluded because of image quality. We compared total grey matter volume across participants. There were no outliers regarding total grey matter volume compared to mean grey matter volume. After final visual inspection, smoothed normalized grey matter maps from all participants were included for statistical analysis.

### 2.6 VBM region-of-interest

We conducted two region of interest (ROI) analyses to investigate the association between apathy and VS as well as apathy and DS volume. We used the same functionally defined VS mask from our previous fMRI studies (Kirschner et al., 2019; Kirschner et al., 2016; Kirschner et al., 2018; Kluge et al., 2018) to allow direct comparison between structural and functional findings (left VS: x,y,z=-12,10,-2, MNI coordinates from Knutson and Greer) (Fig. 3A). This approach is similar to Roth and colleagues (Roth et al., 2016) who also used a VS mask from a previously published fMRI study on VS dysfunction in SZ (Juckel et al., 2006). The ROI was constructed with the Individual Brain Atlases tool in SPM (IBASPM 71) (Alemán-Gómez et al., 2006) implemented in the Wake Forest University Toolbox (Maldjian et al., 2003) (Fig. 3A).To investigate different effects of VS and DS we selected two additional ROIs, which have been used for the same purpose in a recent fMRI study using a subset of the included sample (Stepien et al., 2018). These ROIs were defined by the striatal subregions based on the atlas of Mai and colleagues (Mai et al., 2015) and used previously from Mawlawi and colleagues (Mawlawi et al., 2001) (Fig. 3B) Specifically, the VS mask encompasses the nucleus accumbens, ventral caudate, and ventral putamen, while the DS consists of dorsal caudate and dorsal putamen (Mai et al., 2015).

**Figure 3.**
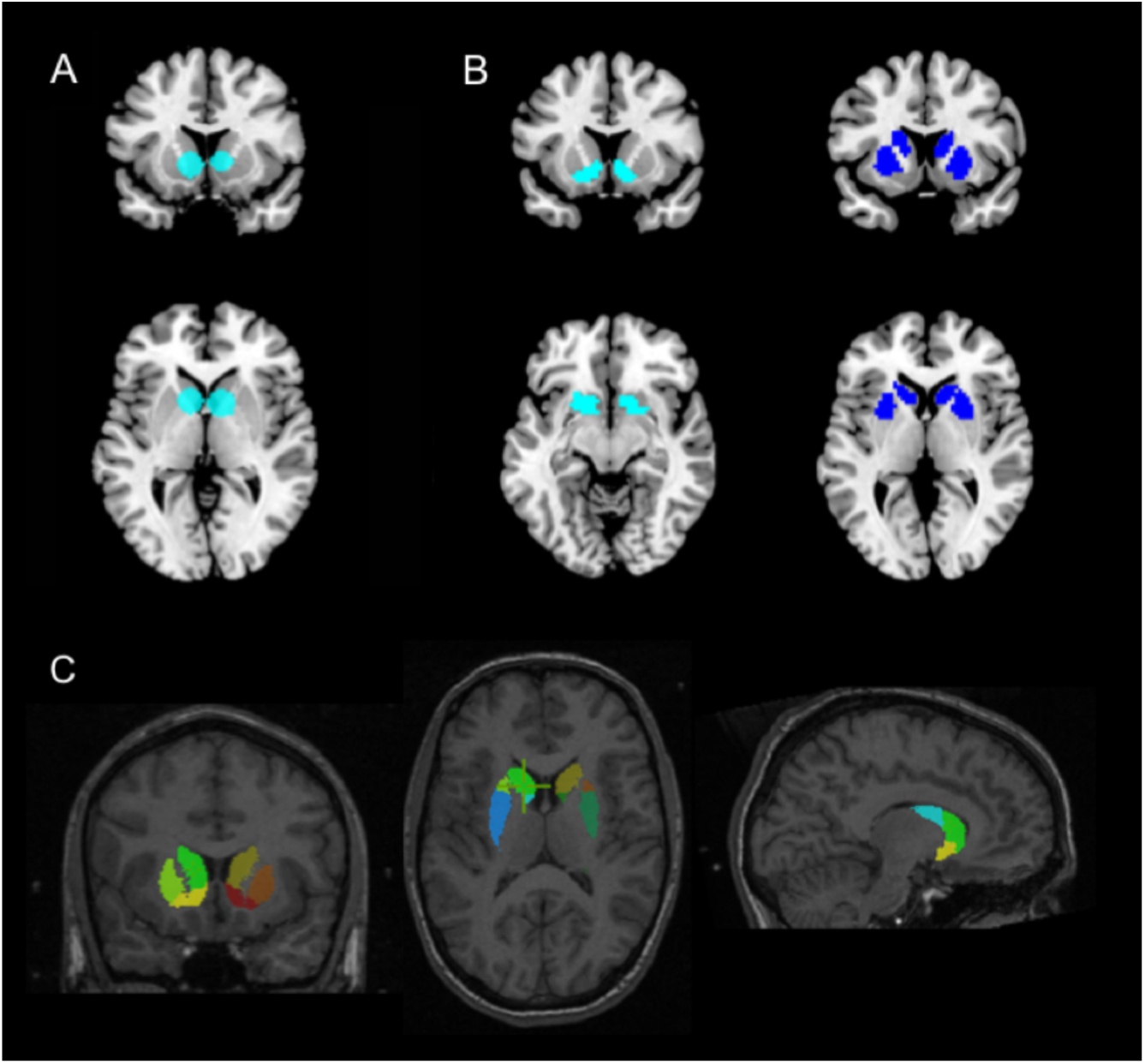
Regions of interest of the VS and DS (A) VS ROI derived from Knutson and Greer (Knutson and Greer, 2008), (B) VS and DS ROI derived from Mawlawi and colleagues (Mawlawi et al., 2001). In total, both ROI approaches results in 4 VS ROIs (2x left, 2xright) and 2 DS ROIs (1xleft, 1xright) (C) Example of how MAGeT delineates the striatal subdivisions on an individual subject scan (Caravaggio et al., 2018a).

### 2.7 VBM data analysis

We analysed the left and right VS and DS (in all 6 ROIs) performing statistical parametric mapping using voxel-wise general linear modeling and random-effects analysis in SPM12. We examined the correlation between apathy scores and VS and DS volume controlling for age, gender and total grey matter volume using one-sample t-test in patients with schizophrenia. To evaluate whether this effect is specific to apathy, this analysis was repeated with the diminished expression scores. We controlled for multiple comparisons using a peak-level family wise error rate of p<0.05 for each ROI separately. In addition, we investigated group differences between patients with schizophrenia and HC using two sample t-test controlling for age, gender and TGMV as covariates (see supplementary Results). Finally, we conducted exploratory whole-brain analyses to investigate group differences (two-sample t-test) and correlations between apathy scores and grey matter volume outside the ROIs (controlling for age, sex and total grey matter volume). We corrected for multiple comparisons using a cluster defining threshold of p<0.001 and a cluster-level family wise error correction of p<0.05 (see supplementary Results).

### 2.8 Multiple Automatically Generated Templates algorithm

Following the analytical approach of Caravaggio and colleagues we extracted subcortical volumes of patients with schizophrenia using MAGeT Brain. The MAGeT-Brain algorithm (Pipitone et al., 2014) was employed to provide fully-automated segmentation of striatal subdivisions (Caravaggio et al., 2018b; Chakravarty et al., 2006).

The delineation of the striatal subdivisions based on serial histological data, and the Collin27 Brain atlas (http://www.bic.mni.mcgill.ca/ServicesAtlases/Colin27), has been described in detail elsewhere (https://github.com/CobraLab/atlases) (Chakravarty et al., 2006). These include the pre-commissural caudate, post-commissural caudate, pre-commissural putamen, post-commissural putamen, and the VS. Briefly, the putamen (and caudate) can be divided based on the position of the anterior commissure as seen on the coronal plane (Fig. 4). The portion of the putamen located anteriorly to the coronal location of the anterior commissure is demarcated as pre-commissural. The remaining portions of the putamen posterior to the anterior commissure, is considered post-commissural (Al-Hakim et al., 2007; Tziortzi et al., 2011). Several studies have been conducted to validate the reliability of MAGeT-Brain against “gold-standard” manual segmentation – the correlation between methods in the striatum have been reported to be around r=0.92 (p=0.0001) with a DiceKappa of 0.861 (Chakravarty et al., 2013; Makowski et al., 2018). Typically, in a multi-atlas segmentation approach, manually drawn labels from atlases are warped (or propagated) into native subject space by applying transformations estimated from non-linear image registration. Candidate labels from all atlas images are fused via probabilistic segmentation techniques to create a final segmentation. The goal of the MAGeT-Brain algorithm is to mitigate sources of error from regular multi-atlas segmentation approaches, such as: 1) spurious non-linear registration or resampling errors (e.g. partial volume effects in label resampling), and 2) irreconcilable differences in neuroanatomy between the atlas and target images. The MAGeT-Brain algorithm is a modified multi-atlas segmentation technique, which employs a limited number of high-quality manually segmented atlases as an input to reduce bias and enhance segmentation accuracy. MAGeT-Brain propagates atlas segmentations to a template library, formed from a subset of target images, via transformations estimated by nonlinear image registration. The resulting segmentations are then propagated to each target image and fused using a label fusion method. Randomly selected cases (n=21) were selected to create a template library through which the final segmentation was bootstrapped. Each subject in the template library was segmented through non-linear atlas-to-template registration followed by label propagation, yielding a unique definition of the subdivisions for each of the templates. The bootstrapping of the final segmentations through the template library produces candidate labels for each subject, and the labels are then fused using a majority vote to complete the segmentation process. Since this is a majority vote process, to avoid potential “ties”, an odd number of template images were employed. Non-linear registration was performed using a version of the Automatic Normalization Tools (ANTS) registration technique (Avants et al., 2008) that is compatible with the minc toolkit (https://github.com/vfonov/mincANTS). Volumes (mm^3^) from ROIs were averaged across hemispheres. Quality control by visual inspection was carried out by author FC to ensure that, there were no major artifacts in the original T1 images and no anomalies in the labelling of the subcortical structures by examining for each subject the original subject image with the resulting labelled image. Total brain volume (TBV) was obtained using the Brain Extraction based on non-local Segmentation Technique (BEaST) method (Eskildsen et al., 2012). BEaST is designed to include CSF (in the ventricles, cerebellar cistern, deep sulci, along surface of brain, and brainstem), the brainstem, and cerebellar white matter (WM) and grey matter (GM) in the brain mask, while excluding the skull, skin, fat, muscles, dura, eyes, bone, exterior blood vessels, and exterior nerves.

**Figure 4.**
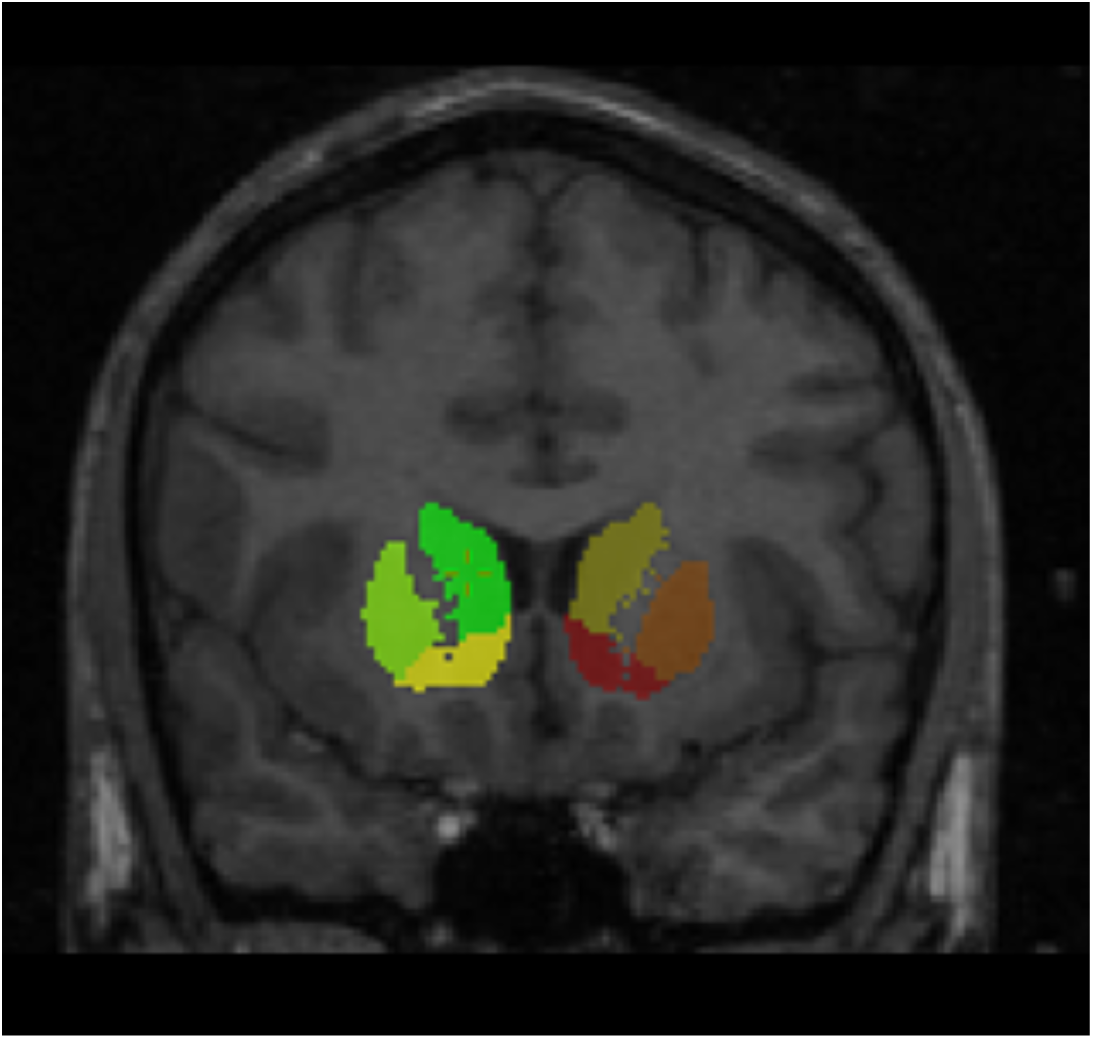
Coronal plane illustrating how the putamen (and caudate) can be divided based on the position of the anterior commissure.

### 2.9 MAGeT-Brain data analysis

We used non-parametric Spearman partial correlations to investigate the relationship between volumes of left and right VS volume, derived from the MAGeT-Brain algorithm, and apathy scores controlling for age, gender and TBV (SPSS Statistics 25, IBM). To identify potential effects specific to apathy and not global negative symptoms we repeated the analysis with diminished expression scores. To address potential confounds, we correlated the extracted left and right VS volumes with age of onset, duration of illness and current antipsychotic medication (measured in chlorpromazine equivalence (CPZ)).

### 2.10 Correlation analysis of VS fMRI activity and VS volume

Given that reduced left VS activation during reward anticipation is associated with apathy (Fig. 2), we examined whether left VS activity is also related to left VS volume. fMRI data from the left VS ROI, x,y,z=-12,10,-2) (Knutson and Greer, 2008) were available from a subset of n=55 patients with schizophrenia. Using Spearman correlations, we assessed the relationship between this individual mean contrast estimate of high reward anticipation versus no reward anticipation and left VS volume (derived from the MAGeT-Brain algorithm) controlling for age, gender and TBV.

## 3. Results

### 3.1 Demographics

Participant characteristics, clinical data and group comparisons are summarized in Table 1. Patients with schizophrenia and healthy controls did not significantly differ in age (SZ, mean=30.5 (8.4); HC, mean=30.3 (8.2), t=0.168, p=0.87) and gender (SZ, n=45 male/15 female, HC, n=34 male/24 female, X^2^=3.576, p=0.06). None of the BNSS negative symptom scores correlated with positive symptoms, measured with the PANSS positive factor (BNSS apathy: r_s_=-0.01, p=0.94; BNSS diminished expression, r_s_ =-0.15, p=0.24), depression measured with the CDSS (BNSS apathy: r_s_=0.15, p=0.25; BNSS diminished expression, r_s_=0.13, p=0.32), and current medication dose measured with chlorpromazine equivalents (BNSS apathy: r_s_=-0.05, p=0.68; BNSS diminished expression, r_s_=0.17, p=0.19). This suggests that negative symptoms were not secondary due to other symptom dimensions or medication (Kirschner et al., 2017).

**Table 1.** Demographic, Psychopathological and Clinical Data

|  | HC (n = 58) | SZ (n = 60) | Statistical test | p value |
| --- | --- | --- | --- | --- |
| Age, years | 30.3 (8.2) | 30.5 (8.4) | $t = 0.168$ | 0.87 |
| Gender m; f (n, %) | 34 m; 59% / 24 f; 41% | 45 m; 75% / 15 f; 25% | $\chi^2 = 3.576$ | 0.06 |
| Education, years | 14.3 (2.5) | 11.8 (2.5) | $U = 791.5$ | < 0.001 |
| Age of onset, years |  | 22.2 (5.4) |  |  |
| Duration of illness, years |  | 8.4 (7.3) |  |  |
| CPZ equivalents |  | 494.5 (456.7) |  |  |
| BNSS Total |  | 24.1 (13.0) |  |  |
| BNSS Apathy |  | 15.5 (8.5) |  |  |
| BNSS Diminished Expression |  | 8.0 (7.3) |  |  |
| PANSS Positive Factor |  | 6.6 (2.5) |  |  |
| PANSS Negative Factor |  | 13.2 (5.6) |  |  |
| PANSS Depressive Factor |  | 5.5 (2.2) |  |  |
| PANSS Total |  | 48.7 (10.4) |  |  |
| CDSS |  | 1.8 (2.2) |  |  |
| GAF |  | 57.6 (9.7) (n=47) |  |  |

*Note: Data are presented as means and standard deviations. BNSS, Brief Negative Symptom Scale; PANSS, Positive and Negative Syndrome Scale; Diagnosis; BNSS Apathy = Avolition, Anhedonia, Asociality Dimension; BNSS Diminished Expression = Affective Flattening or Blunting, Alogia Dimension. We investigated potential group differences using 2-sample $t$ tests for continuous data and $\chi^2$ tests for categorical data. For non-normal distribution data, we applied Mann-Whitney-U-tests.*

### 3.2 Correlation between apathy and striatal Volume

The first aim of the study was to replicate and extend previous work reporting an association between VS volume and apathy in patients with schizophrenia. Using VBM, we did not find a significant correlation between VS or DS grey matter volume and apathy scores controlling for age, gender and TGMV (SVC, peak-level p_FWE_<0.05). Diminished expression scores were not correlated with VS or DS grey matter volume (SVC, peak-level p_FWE_<0.05). Similarly, using the MAGeT-Brain algorithm, apathy did not correlate with left or right VS volume controlling for age, gender and TBV (left: r_s_=0.09, p=0.49; right: r_s_=0.017, p=0.9). Likewise, diminished expression scores did not correlate with either left or right VS volume (left: r_s_=0.072, p=0.59; right: r_s_=0.074, p=0.59). Taken together, we failed to replicate previous findings applying both previously used analytical approaches.

### 3.3 Correlation between potential confounds and VS volume

To test whether other clinical and demographic characteristics are related to VS volume, we correlated age, age of onset, duration of illness and current antipsychotic medication (CPZ) with VS volume derived from the MAGeT-Brain analysis. Partial Spearman correlations revealed no significant effect of age (left: r_s_=-0.101, p=0.44; right: r_s_=-0.158, p=0.23), age of onset (left: r_s_=0.133, p=0.32; right: r_s_=0.114, p=0.40), duration of illness (left: r_s_=-0.133, p=0.44; right: r_s_=-0.103, p=0.44) and current antipsychotic medication (CPZ) (left: r_s_=-0.098, p=0.47; right: r_s_=-0.089, p=0.51).

### 3.4 Reduced left VS volume is associated with left VS hypoactivation

Next, we asked whether left VS volume is associated with left VS BOLD activation during reward anticipation, in schizophrenia patients showing a relationship between blunted left VS activation and apathy (Fig. 2). Using Spearman correlation, we found a trend-level correlation between left VS volume and left VS BOLD activation during reward anticipation in patients with schizophrenia (r_s_=0.248, p=0.067) (Fig. 5). Calculating the coefficient of determination (r^2^=0.062) from the correlation coefficient (r_s_=0.248) revealed that left VS volume explained 6% of the variance in left VS BOLD activity during reward anticipation. Controlling for age, gender and TBV, this correlation remained at the same effect size (r_s_=0.244, p=0.081, r^2^=0.058). Taken together, we observed a small to moderate relationship between left VS volume and VS BOLD activity during reward anticipation in SZ patients.

**Figure 5.**
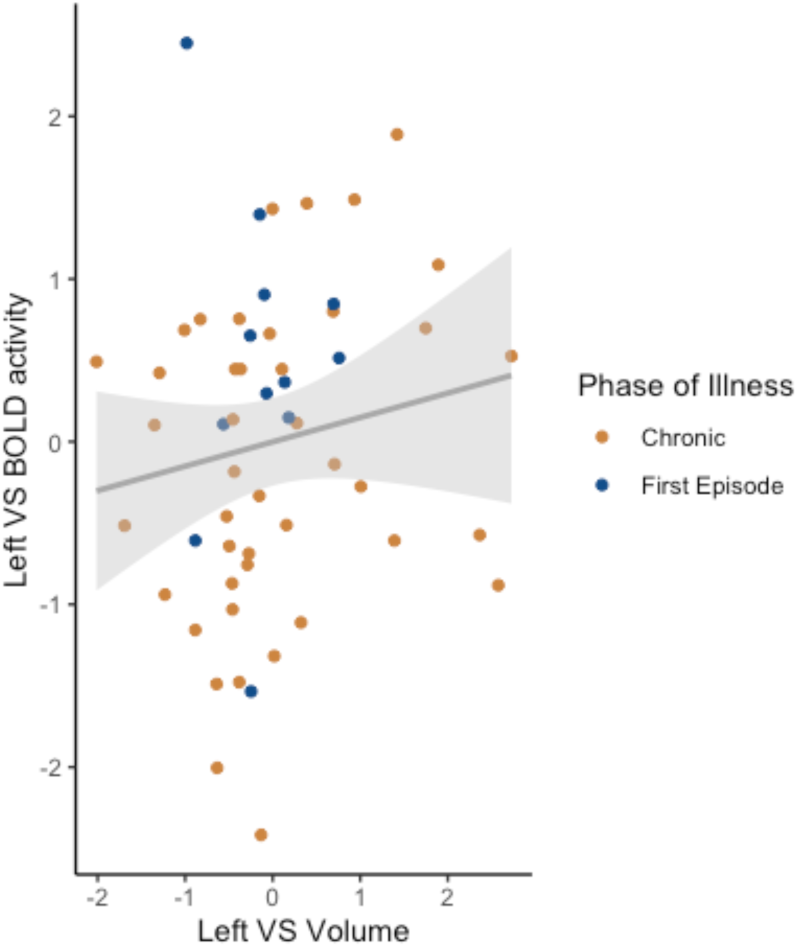
Correlation between left VS volume and left VS BOLD activity during reward anticipation in patients with schizophrenia. Reduced left VS volume was associated with reduced left VS BOLD activity during reward anticipation (r_s_=0.248, p=0.067).

## 4. Discussion

Capitalizing on a sample of SZ patients exhibiting blunted VS BOLD activation related to apathy, the present study aimed to extend previous work on the relationship between structural VS abnormalities and the severity of apathy in schizophrenia. Although applying the same methodological approaches as two recently published studies (Caravaggio et al., 2018a; Roth et al., 2016), we could not replicate the previously observed association between apathy and reduced VS volume in a larger sample. Given the fact that we have found blunted VS activation related to apathy in the same dataset (Fig. 2), the present findings suggest that functional and structural striatal correlates of apathy can occur separately. Compared to apathy, other clinical (e.g. duration of illness, age of onset, medication) and demographic characteristics (e.g. age) might be more strongly related to striatal volume change, possibly limiting the ability to replicate previous findings. Interestingly, and in contrast to our negative findings regarding a relation between apathy and VS volume, we observed a small to moderate correlation between reduced left VS volume and blunted left VS BOLD activation during reward anticipation. This suggests an independent functional-structural relationship in the VS, which could be related to more general disease processes and not directly to apathy.

The negative findings of the present study raise the question whether differences in sample characteristics (compared to (Caravaggio et al., 2018a; Roth et al., 2016)) are able to explain the missing association between reduced VS volume and apathy. Relevant confounding factors are differences in age, duration of illness and antipsychotic medication dose (Amato et al., 2017; Haijma et al., 2013; Hashimoto et al., 2018; van Erp et al., 2016). Compared to the patient group in the current study (n=60, age, mean=30.5, SD=8.4), patients in the study from Roth et al. were on average almost 10 years older (n=23, age, mean=39.4, SD=12.3) and on average around 30 years older in the study from Caravaggio et al. (n=41, age, mean=60.2, SD=6.7) (Caravaggio et al., 2018a; Roth et al., 2016). Accordingly, in the latter study, patient’s illness duration was on average over 25 years longer and apathy/amotivation correlated positively with age (Caravaggio et al., 2018a). In a recent ENIGMA meta-analysis van Erp. and colleagues showed a positive association between duration of illness and age with putamen and pallidum volume (van Erp et al., 2016). This is in contrast to converging findings from studies in healthy individuals, showing a constant decline in subcortical volumes with age (Goodro et al., 2012; Jernigan et al., 2001; Walhovd et al., 2005; Wang et al., 2019). One likely explanation for these divergent findings is that the association between illness duration and subcortical volumes may partly be impacted by cumulative antipsychotic drug exposure (van Erp et al., 2016). Accumulating evidence from human and animal studies shows that long-term antipsychotic medication leads to subcortical grey matter enlargement (Amato et al., 2017). Even short-term use of antipsychotic medication in first-episode psychosis led to enlargement of the right globus pallidus (Albacete et al., 2019). Longitudinal studies in medication-naïve and minimal-treated patients observed increases of putamen and caudate volume after antipsychotic treatment (Glenthoj et al., 2007; Ho et al., 2011; Li et al., 2012). Correspondingly, two meta-analyses on cross-sectional data reported a positive association between caudate volume (Haijma et al., 2013) and pallidum volume (Hashimoto et al., 2018) with current medication dose at time of scanning. However, the cross-sectional design of the present study and missing information on medication history or long-term cumulative antipsychotic dose limited the ability to further investigate the potential interaction between apathy, antipsychotic medication and subcortical abnormalities.

Different subtypes of negative symptoms might contribute to the observed negative findings. In this study, patients were in an earlier stage of disease and were on a stable medication dose for a minimum of two weeks, while one third of patients received more than one antipsychotic medication (Supplementary Table S1). It is possible that, in some patients, negative symptoms were transient and did not become persistent or enduring after a clinical stabilization of minimum six months (Buchanan, 2007; Hovington and Lepage, 2012). In contrast, Caravaggio and colleagues (Caravaggio et al., 2018a) included only stable patients with no inpatient hospitalization in the last six months and a stable dose for at least six months of either olanzapine or risperidone monotherapy. Based on the criteria proposed by Buchanan (Buchanan, 2007), we can consider negative symptoms of those patients as persistent. Although not fulfilling the criteria for persistent negative symptoms, Roth and colleagues (Roth et al., 2016) included only patients from outpatient clinics on stable medication for at least three months. This suggests a similar pattern of more persistent negative symptoms compared to the present study. Regarding structural brain abnormalities, studies found preliminary evidence for an association between persistent negative symptoms and grey matter reduction within regions of the fronto-limbic circuit (Hovington and Lepage, 2012). However, the association with specific basal ganglia volume abnormalities remains to be determined. The distinction between primary and secondary negative symptoms might also be relevant for the inconclusive findings. In this study, negative symptoms were not related to secondary sources such as medication, depression or positive symptoms suggesting that patients had primary negative symptoms (Carpenter et al., 1985; Kirschner et al., 2017). In contrast, Caravaggio and colleagues found a positive correlation between apathy/amotivation and positive symptoms, which suggested that negative symptoms were at least partly caused by persistent positive symptoms (Carpenter et al., 1985; Kirschner et al., 2017). Taken together, different clinical manifestations of transient vs. persistent and primary vs. secondary negative symptoms could contribute to the divergent findings of striatal abnormalities.

Although VS volume did not correlate with apathy, we found a small trend-level association between left VS volume and blunted VS BOLD activation during reward anticipation. This finding is consistent with previous ones in patients with bipolar disorder. Caseras and colleagues reported an association between left putamen volume and left VS activation during reward anticipation (Caseras et al., 2013), while Yip and colleagues found a relationship between right putamen volume and right DS activation during reward anticipation (Yip et al., 2015). However, the relationship between striatal volume and function was not observed with working-memory related BOLD activation in schizophrenia (Ehrlich et al., 2012). This suggests a potential specific association between striatal volume and reward-related activation. Future work should confirm our preliminary findings and elucidate whether this structural-functional relationship in the VS is a trans-diagnostic mechanism of the schizophrenia-spectrum that is not related to apathy.

### 4.1 Limitations and future directions

Although we took advantage of a cohort with well-characterized negative symptoms and controlled for several factors such as secondary negative symptoms and current daily medication dose, this study has some limitations inherent to cross-sectional designs. First, retrospective information on medication history were limited and are prone to inaccuracy. Therefore, we could not address the long-term effects of cumulative antipsychotic exposure and potential divergent effects of different medication types (Albacete et al., 2019; Ebdrup et al., 2013). Second, negative symptoms vary over time and long-term clinical records would be favorable to identify subtypes of negative symptoms. As a consequence, future longitudinal studies are needed to examine the interactions between long-term antipsychotic exposure, variation in negative symptoms and duration of illness to clarify the role of striatal structural abnormalities in the pathophysiology of apathy. In addition, multimodal imaging approaches should further explore the relationship between VS structure and function in various task conditions and at rest. To this end, these studies should not be limited to chronic schizophrenia but should expand across the complete schizophrenia-spectrum. This will help to elucidate trans-diagnostic neural mechanisms underlying the pathophysiology of apathy and motivational deficits.

### 4.2 Conclusions

Taken together, by leveraging a larger patient sample with established striatal functional abnormalities and apathy, we failed to replicate previous reports of an association between reduced VS volume and apathy. These findings provide evidence that functional and structural correlates of apathy can occur independently and suggest a stronger relationship between VS function and apathy compared to VS structure and apathy.

## Data Availability

Full Data are available on request.

## Contributors

A. Burrer, M. Kirschner, S. Kaiser, E. Seifritz and F. Caravaggio designed the study. M. Kirschner, A. Manoliu, K. Gütter, P. Stämpfli acquired the MRI data. A. Burrer, A. Manoliu, A. Schmidt, S. Borgwardt and B. Habermeyer conducted the VBM analysis. F. Caravaggio, E. Plitman, M. Chakravarty and A. Graff-Guerrero conducted the MAGeT Brain analysis. M. Lepage, A. Dagher and A. Abivardi contributed to the statistical analyses. A. Burrer and M. Kirschner wrote the first draft of the manuscript. All authors revised the manuscript. All authors contributed to and have approved the final manuscript.

## Acknowledgment

We want to express our gratitude to the participants of the study.

**Appendix: Supplementary data**

